# The effectiveness of digital technology interventions to reduce loneliness in adult people: A protocol for a systematic review and meta-analysis

**DOI:** 10.1101/19000414

**Authors:** Syed Ghulam Sarwar Shah, David Nogueras, Hugo Cornelis van Woerden, Vasiliki Kiparoglou

## Abstract

**Introduction:** Loneliness is an emerging public health problem, which is associated with social, emotional, mental and physical health issues. The application of digital technology (DT) interventions to reduce loneliness has increased in recent years. The effectiveness of DT interventions needs to be assessed systematically.

**Methods and analysis:** *Aim:* To undertake a systematic review and meta-analysis on the effectiveness of digital technology interventions to reduce loneliness among adult people.

*Design:* systematic review and meta-analysis.

*Data sources:* Five leading online bibliographic databases: PubMed, Medline, CINAHL, EMBASE, and Web of Science.

*Publication period:* 1 January 2010 to 30 April 2019.

*Inclusion criteria:* Primary studies involving the application of digital technology interventions to reduce loneliness, involving adult participants (aged 18 years and more) and published in the English language.

*Search strategy:* Literature searches using a priory list of keywords, involvement of two independent researchers in article screening, short listing and data extraction using a predefined template based on the population, intervention(s), comparator(s) and outcome(s) (PICO) framework.

*Synthesis and meta-analysis:* A narrative summary of the characteristics of included studies, findings by the type of DT intervention, and the age, gender and ethnicity of participants. A meta-analysis by the type of DT intervention and determination of effect sizes.

*Quality of evidence and bias:* Quality of evidence assessed the RoB 2.0 (revised tool for Risk of Bias in randomized trials) and ROBINS-I (Risk Of Bias in Non-randomized Studies - of Interventions) tools for randomized control trials and non-randomized studies respectively. Heterogeneity between studies determined by the I^2^ and Cochran’s Q statistics and publication bias checked with funnel plots and the Egger’s test.

*Patients and public involvement:* None

*Ethics and dissemination:* Ethics approval was not required for writing this protocol. The findings will be disseminated through the publication of research articles and conference presentations.

*PROSPERO Registration Number:* CRD42019131524.

**Article Summary:** *Strengths and limitations of this study:*

- The main strength of this study includes a systematic assessment of evidence on the effectiveness of digital technology interventions to reduce loneliness, which is imperative from the health and social care and public health perspectives.
- Another strength of the study is the involvement of two independent researchers (and a third researcher as an arbitrator) involved in the identification, screening, inclusion and extraction of on a predefined template using the PICO framework.
- Limitations may include missing identification of additional relevant studies due to the application of selection filters such as the publication years and English as the publication language.

## INTRODUCTION

### Definitions

Loneliness is defined as a feeling of distress which develops in an individual when the social relationship is perceived as less satisfying than the desired level.^[1]^ Loneliness refers to a gap between the desired and the actual social relationships of an individual.^[2]^ Loneliness and social isolation are often reported together in the literature; however, there is a difference between these two terms. Loneliness is a subjective feeling while social isolation is an objective feeling and the former arises because of the perceived gap between the actual and the desired social relationships while the latter develops due to the absence of social contact with the society, individuals,^[2]^ family and friends.^[3]^ Nonetheless, loneliness and social isolation are different from each other^[3]^ with distinct pathways to adverse health effects.^[4,5]^ Tackling both loneliness and social isolation is important because they are associated with adverse health effects.^[3]^

### Burden of loneliness

Loneliness is increasing, especially in the developed countries^[6]^ such as Australia,^[7]^ Japan,^[8]^ the UK^[9]^ and the US.^[10]^ Loneliness is an epidemic^[5]^ and a rising public health problem^[11]^ because it leads to social, mental and health problems.^[12–14]^

#### Loneliness and demographic factors

Loneliness can affect individuals of any age and members of any community.^[2]^ It is prevalent in young children and adolescents^[15]^ who are more susceptible to loneliness as they go through social and personal transformations.^[16]^ In children and adolescents, loneliness is associated with chronic physical conditions.^[17]^ In older adults, loneliness is statistically significantly associated with female gender, older age, poor income, lower educational level, living alone, low quality of social relationships, poor self-reported health, poor functional status, marital status as unmarried^[18]^ including people who have never been married, are widowed or divorced.^[19]^ In older people, loneliness is common because they are more vulnerable because of age-related changes and losses.^[20]^

#### Loneliness and physical and mental health

Loneliness is associated with health and mental health risks.^[20]^ Loneliness is also positively associated with chronic conditions such as cancer^[19]^ and cardiovascular disease^[21]^ and it has a negative influence on the quality of life, health and survival.^[22]^ In old age, loneliness is associated with reduced quality of life, poor health, maladaptive behaviour, and depressed mood^[23]^ as well as increased risk of developing Alzheimer’s disease.^[24]^

Loneliness is associated with mental health such as depression, low self-esteem, anxiety, perceived stress^[16,25]^, psychosis,^[26]^ shame and fear,^[18]^ incident dementia,^[27]^ sleep disorders such as severe insomnia^[28]^ and suicidal ideation in older adults.^[29]^

Thus, loneliness increases risks not only to the physical health^[5,30]^ and mental health^[12–14]^ but also increases risk of early/premature mortality and all-cause mortality^[5]^ especially in older men and women.^[31]^

#### Loneliness and social and environmental factors

Loneliness is also associated with social and environmental factors such as boredom and inactivity, the role of the recent losses of family and friends, inaccessible housing, inadequate resources for socialising, unsafe neighbourhoods, migration patterns and environmental barriers.^[18]^ In addition, loneliness has a positive association with lack of psychological or social support,^[19]^ low economic level and living arrangements.^[32]^

More importantly, not only adults at the risk of loneliness are increasing but at the same time costs associated with loneliness are also increasing.^[33]^ It is therefore imperative to tackle this double edge sword like situation created by loneliness through a range of interventions and strategies.

### Interventions to tackle loneliness

Tackling loneliness requires improvement of social skills, increase in social support, enhancement of social contact opportunities and tackling the maladaptive social cognition.^[33]^ Intervention applied to reduce loneliness could be broadly divided into two categories i.e. social interventions and technological interventions.

Social interventions applied to reduce loneliness include befriending, residential and school-based camps, reminiscence therapy, animal interventions, gardening, physical activity, and technology of many kinds.^[34]^ In addition, interventions focusing on social network maintenance and enhancement have also been applied and found useful to combat loneliness.^[20]^ Social interventions could also be combined with the application of the technology such as online peer-to-peer interactions and support groups through social media platforms to alleviate loneliness especially in persons with psychotic disorders.^[35]^ However, loneliness in older people can create serious problems that could not be alleviated with the social support only^[32]^ but it requires other types of interventions such as technological interventions (e.g. digital applications (apps), online social networks and social robots), which could enhance perceived emotional support and social interaction and thereby help in promoting health in older adults.^[36]^

#### Digital technology interventions

The term ‘digital technology’ (DT) refers to the technology, equipment and applications that process information in the form of numeric codes, usually a binary code, which is processed and used by many devices such as computers, smartphones and robots.^[37]^ Research in DT has become imperative for various reasons such as rapid development in DTs, social change and ubiquity of computer technologies, which are an integral part of the daily life of many people.^[38]^ There are many kinds of technology interventions that could be applied to reduce loneliness.^[34]^

We therefore focus on DT interventions to reduce loneliness. We will assess any intervention that involves the application of DT to reduce and alleviate loneliness in the adult population.

### Previous systematic reviews on technological interventions for loneliness

There are some systematic reviews on technological interventions for tackling loneliness. For example, Pearce et al. (2012) undertook a systematic review on the availability and use of robotics by older people and reported that robotic technologies could support older people and those with disabilities in independent living; however, they suggested that there was a need for further research to achieve the full potential of robotic technologies on social connectedness.^[39]^ In addition, a meta-analysis on the effectiveness of computer and Internet training interventions intended to reduce loneliness and depression in older adults by Choi et al. (2012) reported that computer and Internet programs were effective in managing loneliness among older adults.^[40]^

Morris et al. (2013) conducted a systematic review of literature on the effectiveness of smart-home technologies i.e. passive sensors, monitoring devices, robotics and environmental control systems for promoting independence, health, well-being and quality of life, in older adults and found only one study on the effectiveness of smart-home technologies and they suggested further research in this domain.^[41]^ A literature review of Hagan et al. (2014) investigated the effectiveness of social therapeutic interventions to reduce loneliness in older people and included three studies on new technologies and one on a group work intervention and identified significant reductions in loneliness.^[42]^ Miler et al. (2014) report a systematic review on the effectiveness of virtual reality and gaming system on enabling physical activity in older peoples living at home and reported a high risk of bias and weakness in the studies included in their review.^[43]^

In addition, a systematic review by Morris et al. (2014) evaluated the effectiveness of smart technologies such as tailored internet programs to improve or maintain the social connectedness of older people living at home and found that digital/smart technological interventions have positive outcomes compared to traditional social care interventions.^[44]^ In addition, Morris et al. focused on the effects of smart technologies, which they reported as computers and the internet, on improving or maintaining social connectedness in older people living in homes and reported that these technologies improved social connectedness in these people.^[44]^

### Rational

Technology can provide opportunities for social connectedness and thus help in reducing loneliness in older adults; however, studies involving technological interventions to alleviate loneliness in frail and institutionalized older adults are limited. ^[45]^

For example, a systematic review of Pearce et al. (2012) focused only on robotic technologies and their effectiveness in independent living by older people and suggested that the effect of robotics on older peoples’ safety and social connectedness needs further research.^[39]^ These findings suggest that assessment of the effect of robotic technologies on loneliness needs to be undertaken.

In addition, a systematic review by Morris et al. (2013) involved assessment of the effectiveness of smart-home technologies: passive sensors, monitoring devices, robotics and environmental control systems for promoting independence, health, well-being and quality of life in older adults; however, this study was limited to finding only one study on the effectiveness of smart-home technologies and their focus was not on loneliness but on independent living in older people.^[41]^

Another systematic review by Morris et al. (2014) on smart technologies for improving or maintaining social connectedness has limitations such as a narrow definition of smart technologies, which they searched as ‘computers’ and the ‘internet’ and combined with assistive technologies. ^[44]^ The other weakness in the study by Morris et al.^[44]^ was their focus on social connectedness and older people living in homes, and they also searched articles published between January 2010 and January 2013. In addition, Morris et al. suggested a need for further research regarding smart technologies for reducing loneliness in older adults.^[44]^

We anticipate the publication of new research studies from 2013 to present and a need to assess the latest research involving digital technologies to reduce loneliness in older people. This is imperative because the technology evolves very rapidly and new technologies (e.g. smart speakers, new sensors, new robot versions, etc.) are being developed and becoming available and adopted very rapidly popular and we need to assess their potential impact on adult people with loneliness.

In addition, evidence of technology-assisted interventions to reduce loneliness and their effects on the social health and well-being of older people is limited.^[23]^ Moreover, there could be the latest research interventions involving newer digital technologies to reduce loneliness in older people that need to be assessed.^[42]^ It is also imperative to identify technological interventions that are effective in reducing loneliness^[23]^ and how technological innovations can be promoted, marketed and implemented to benefit older people with loneliness.^[44]^

## SYSTEMATIC REVIEW REGISTRATION

This protocol was registered with the PROSPERO database (www.crd.york.ac.uk/prospero/), which is an International prospective register of systematic reviews^[46]^ on 10^th^ June 2019 with a registration ID of PROSPERO 2019 CRD42019131524^[47]^ and it can be accessed online at http://www.crd.york.ac.uk/PROSPERO/display_record.php?ID=CRD42019131524

## METHODS AND ANALYSIS

For the development and writing of this protocol, we follow the Joanna Briggs Institute’s guidelines and template for writing a protocol for a systematic review of effectiveness evidence and meta-analysis^[48]^ and the PRISMA-P (Preferred reporting items for systematic review and meta-analysis protocols) 2015 checklist,^[49]^ which is given as appendix I.

### Aims and objectives

The aim and objectives of this study are as follows.

#### Aim

- To undertake a systematic review and meta-analysis on the effectiveness of digital technology interventions to reduce loneliness among adult people.

#### Objectives

- To identify digital technology interventions used to reduce loneliness in adult people.
- To assess the effectiveness of digital technology interventions to reduce loneliness in adult people.

### Review question(s)

#### Main review question

How effective are digital technology interventions to reduce loneliness in adult people?

#### Secondary review question

What digital technology interventions are used for tackling loneliness in adult people?

### Main outcome measure

Loneliness will be the main outcome measure in our study.

#### Timing and effect measures

We will include pre- and post-intervention measurement of loneliness using any of the following loneliness measures: UCLA Loneliness Scale, De Jong Gierveld 6-Item Loneliness Scale, Campaign to End Loneliness Measurement Tool and any other loneliness scale (e.g. Single-item questions, also known as self-rating measures of loneliness). ^[50]^ We will include studies that will have at least three months or more time for follow-up of the outcomes.

#### Inclusion and exclusion criteria

We will include and exclude articles based on a set of inclusion and exclusion criteria respectively (Table 1).

**Table 1.**
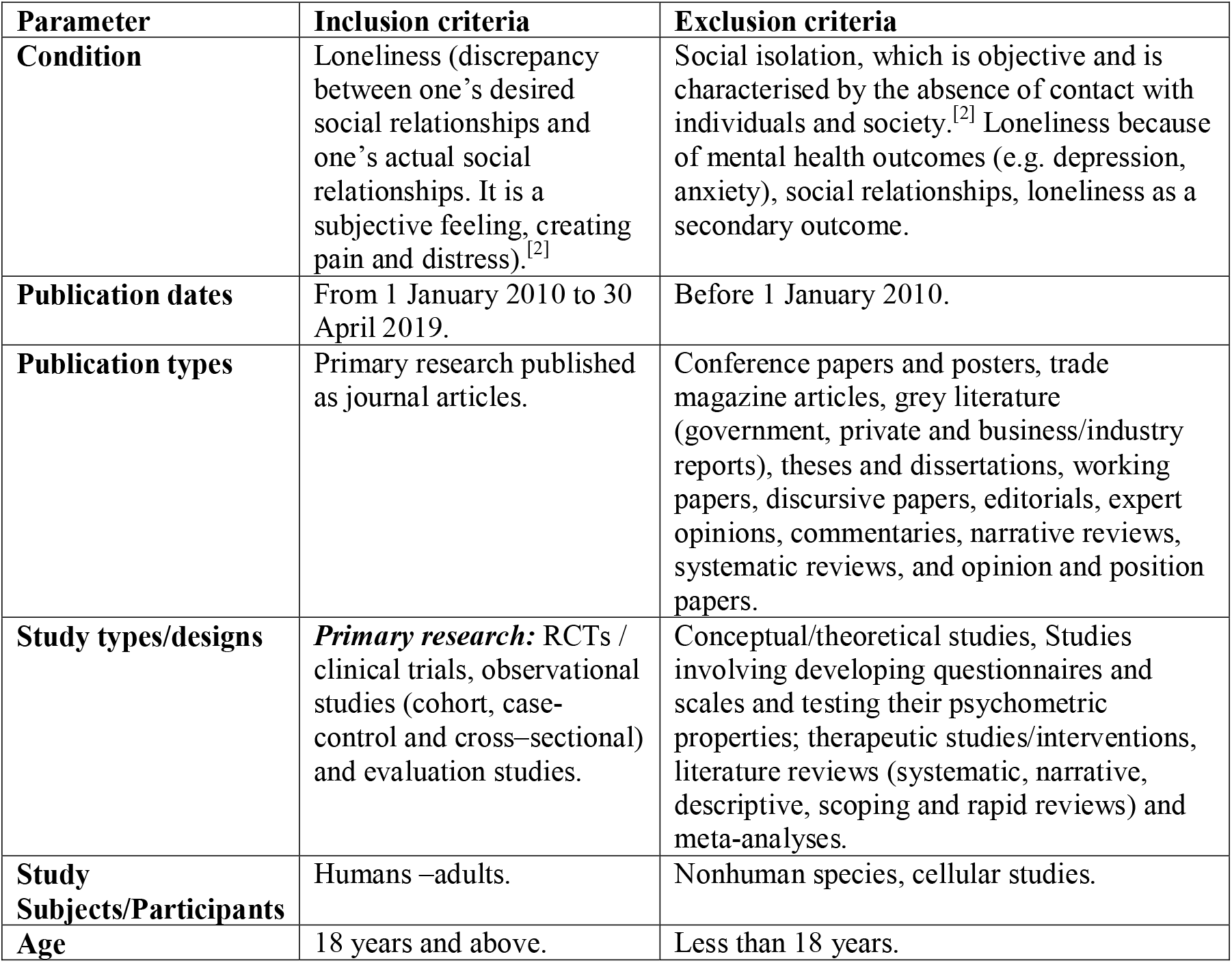
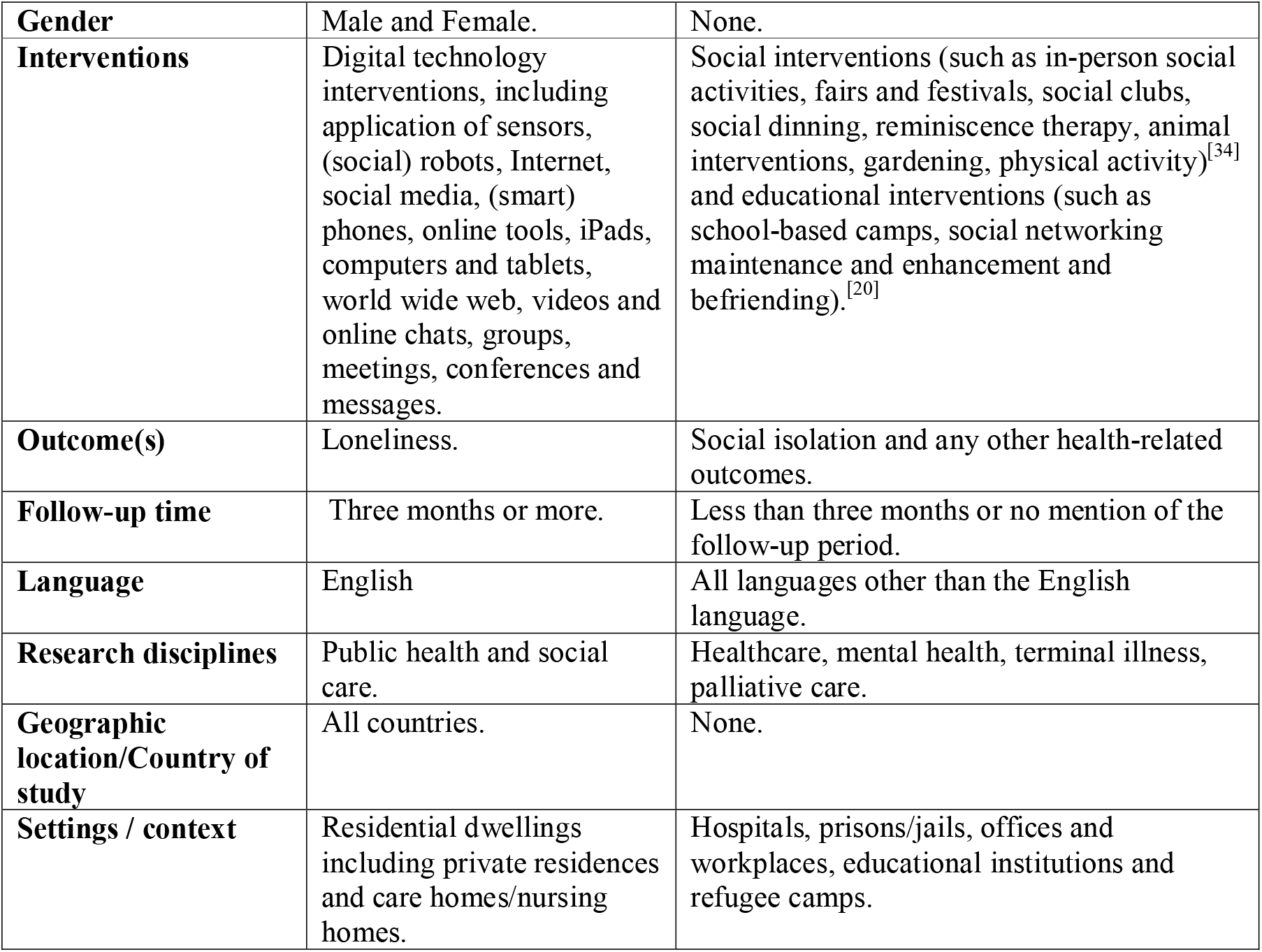
Inclusion and Exclusion Criteria.

| Parameter | Inclusion criteria | Exclusion criteria |
| --- | --- | --- |
| **Condition** | Loneliness (discrepancy between one's desired social relationships and one's actual social relationships. It is a subjective feeling, creating pain and distress).<sup>[2]</sup> | Social isolation, which is objective and is characterised by the absence of contact with individuals and society.<sup>[2]</sup> Loneliness because of mental health outcomes (e.g. depression, anxiety), social relationships, loneliness as a secondary outcome. |
| **Publication dates** | From 1 January 2010 to 30 April 2019. | Before 1 January 2010. |
| **Publication types** | Primary research published as journal articles. | Conference papers and posters, trade magazine articles, grey literature (government, private and business/industry reports), theses and dissertations, working papers, discursive papers, editorials, expert opinions, commentaries, narrative reviews, systematic reviews, and opinion and position papers. |
| **Study types/designs** | **Primary research:** RCTs / clinical trials, observational studies (cohort, case-control and cross-sectional) and evaluation studies. | Conceptual/theoretical studies, Studies involving developing questionnaires and scales and testing their psychometric properties; therapeutic studies/interventions, literature reviews (systematic, narrative, descriptive, scoping and rapid reviews) and meta-analyses. |
| **Study Subjects/Participants** | Humans –adults. | Nonhuman species, cellular studies. |
| **Age** | 18 years and above. | Less than 18 years. |
| **Gender** | Male and Female. | None. |
| **Interventions** | Digital technology interventions, including application of sensors, (social) robots, Internet, social media, (smart) phones, online tools, iPads, computers and tablets, world wide web, videos and online chats, groups, meetings, conferences and messages. | Social interventions (such as in-person social activities, fairs and festivals, social clubs, social dinning, reminiscence therapy, animal interventions, gardening, physical activity)<sup>[34]</sup> and educational interventions (such as school-based camps, social networking maintenance and enhancement and befriending).<sup>[20]</sup> |
| **Outcome(s)** | Loneliness. | Social isolation and any other health-related outcomes. |
| **Follow-up time** | Three months or more. | Less than three months or no mention of the follow-up period. |
| **Language** | English | All languages other than the English language. |
| **Research disciplines** | Public health and social care. | Healthcare, mental health, terminal illness, palliative care. |
| **Geographic location/Country of study** | All countries. | None. |
| **Settings / context** | Residential dwellings including private residences and care homes/nursing homes. | Hospitals, prisons/jails, offices and workplaces, educational institutions and refugee camps. |

### Data sources

We will systematically search for articles published in five large and widely used online bibliographic databases: PubMed, CINAHL, Web of Science, Medline and EMBASE (Excerpta Medica dataBASE), which cover literature in the fields of health sciences, medicine, nursing, allied health, biomedicine, health technology, healthcare, life sciences and biomedical sciences. Literature searches through these five databases will cover publication period from 1 January 2010 to 30 April 2019.

We will also review the references’ lists of shortlisted articles for identifying any relevant studies. We will write to the authors for full copies of any articles that could not be accessed or retrieved full via our library.

### Search strategy and parameters

#### Preparation of a list of keywords

##### Preliminary literature searches

First of all, we did a scoping preliminary literature search in the PubMed database using a set of few keywords, as shown in the search user query shown in Box 1.

###### User Query

Loneliness[MeSH Major Topic] AND needs AND intervention* AND ( ( systematic[sb] OR Review[ptyp] OR Randomized Controlled Trial[ptyp] OR Observational Study[ptyp] OR Meta-Analysis[ptyp] OR Journal Article[ptyp] OR Evaluation Studies[ptyp] OR Clinical Trial[ptyp] ) AND ( “2010/01/01”[PDat] : “2018/11/12”[PDat] ) AND Humans[Mesh] AND English[lang] AND ( Female[MeSH Terms] OR Male[MeSH Terms] ) AND ( jsubsetn[text] OR medline[sb] ) AND ( aged[MeSH] OR middle age[MeSH] OR adult[MeSH:noexp] OR adult[MeSH] OR adolescent[MeSH] OR child[MeSH:noexp] ) )

###### Filters activated

Systematic Reviews, Review, Randomized Controlled Trial, Observational Study, Meta-Analysis, Journal Article, Evaluation Studies, Clinical Trial, Publication date from 2010/01/01 to 2018/11/12, Humans, English, Female, Male, Nursing journals, MEDLINE, Aged: 65+ years, Middle Aged: 45-64 years, Adult: 19-44 years, Adult: 19+ years, Adolescent: 13-18 years, Child: 6-12 years, Field: Title/Abstract

As a result of our preliminary search query, we captured 100 articles, which were reviewed independently by two reviewers (SGSS and DN), who screened titles of all these articles and recommended relevant articles to be included in the second stage of screening i.e. reading the abstracts.

##### List of keywords

We noted subject terms reported in all articles that the two reviewers recommended for the second stage. We reviewed these keywords and prepared a refined list of keywords. We noted that there were very few keywords about various types of digital tools and technologies that we thought could be relevant for identifying digital interventions for addressing the issue of loneliness. Therefore, we divided keywords into two categories, i.e. conditions/issues and intervention / technology (Table 2). A full list of keywords is shown in Table 2 and we will use these keywords for our full literature searches and identifying relevant articles. We will search for these keywords in the titles, abstracts and author keywords fields in the selected databases.

**Table 2.**
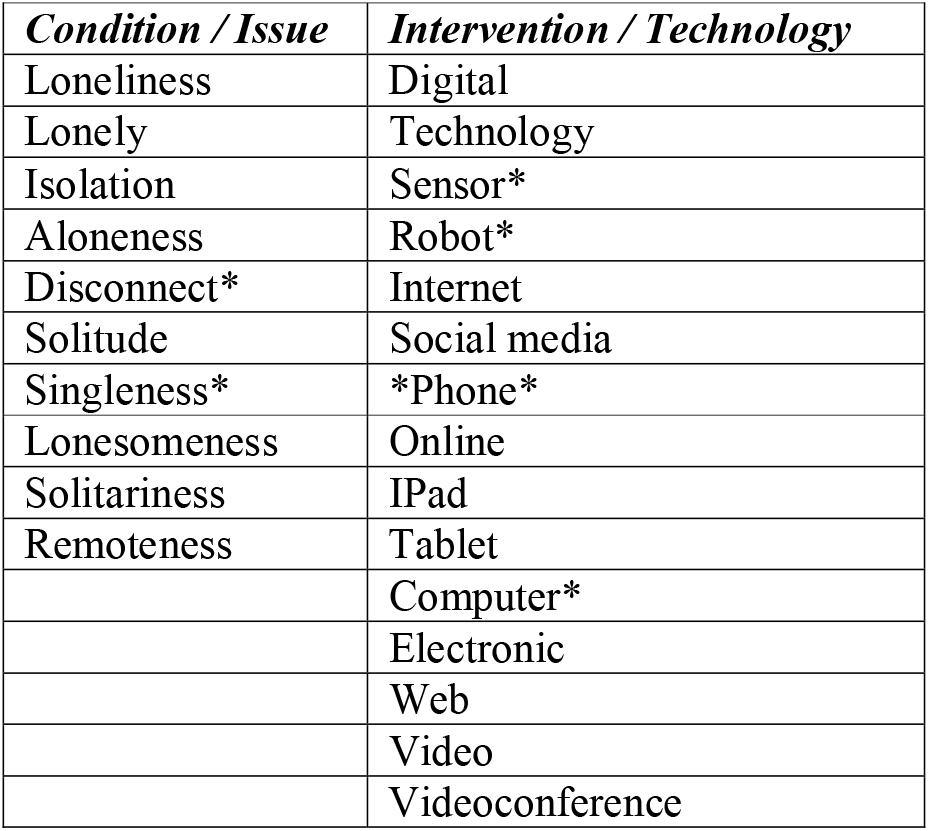
List of keywords.

| ***Condition / Issue*** | ***Intervention / Technology*** |
| --- | --- |
| Loneliness | Digital |
| Lonely | Technology |
| Isolation | Sensor* |
| Aloneness | Robot* |
| Disconnect* | Internet |
| Solitude | Social media |
| Singleness* | *Phone* |
| Lonesomeness | Online |
| Solitariness | IPad |
| Remoteness | Tablet |
|  | Computer* |
|  | Electronic |
|  | Web |
|  | Video |
|  | Videoconference |

#### Running of literature searches

Systematic literature searches will be undertaken in the selected five online bibliographic databases using the pre-identified list of keywords.

The literature will be searched using the keywords that will be searched in only the ‘title’ and ‘abstract’ searched fields in the selected bibliographic databases. The keywords will be used firstly in the ‘subject headings’ such as Medical Subject Headings (MeSH) major terms in the PubMed or equivalent in other databases.

The searches will be filtered by applying the inclusion and exclusion criteria. We will identify literature using the keywords and applying the Boolean operators, i.e. ‘OR’, ‘AND’ and ‘NOT’ whilst searching for literature in different selected electronic/online bibliographic databases, as reported in the next section.

In addition to searching through selected online bibliographic databases, we will search for relevant articles through searching references’ lists of all selected articles.

We will seek support from the Bodleian library staff for running of literature searches.

#### Management of study records/references

We will keep a record of amendments in the protocol, if any, using an Excel spreadsheet. All records found in searches through the selected databases will be directly downloaded and exported to the RefWorks software, which is a web-based bibliography and database manager.^[51]^

We will have a quick look at all articles to check whether any information is missing ensuring completeness of bibliographic details. Also, we will note the total number of articles downloaded. Thereafter, we will identify and remove duplicate articles using the RefWorks software as well as manually. We will note the total number of duplicate articles and all duplicate articles will be deleted and the total number of articles remaining will be noted again.

Thereafter, we will create a list of all unique articles in an Excel spreadsheet to facilitate articles screening and shortlisting independently by two researchers (SGSS and DN). In addition, we will use an Excel spreadsheet for extracting data from shortlisted articles on a predefined template (Table 3). For citing articles in our papers and publications, we will use the Zotero software, which is an open-source and free research tool for references management and citations.^[52]^

**Table 3.** Data extraction form.

|  |
| --- |
| Authors (and Year of publication) |
| Country of study |
| Aim/Objectives of the study |
| Research Design |
| Settings |
| Participants (age, gender and ethnicity) |
| Health/Medical condition |
| Sampling method |
| Sample size |
| Attrition rate |
| Research Method(s)/Data collection tool(s) |
| Intervention(s) |
| Comparator(s) |
| Outcome/Results/Findings |
| Authors' Conclusion |
| Quality of study (Reviewers' evaluation/remarks about the Study) |

We will use the PRISMA flow diagram (Figure 1 for the identification, screening, eligibility and inclusion of relevant studies and data extraction,^[53]^ as explained in the following sections.

**Figure 1.**
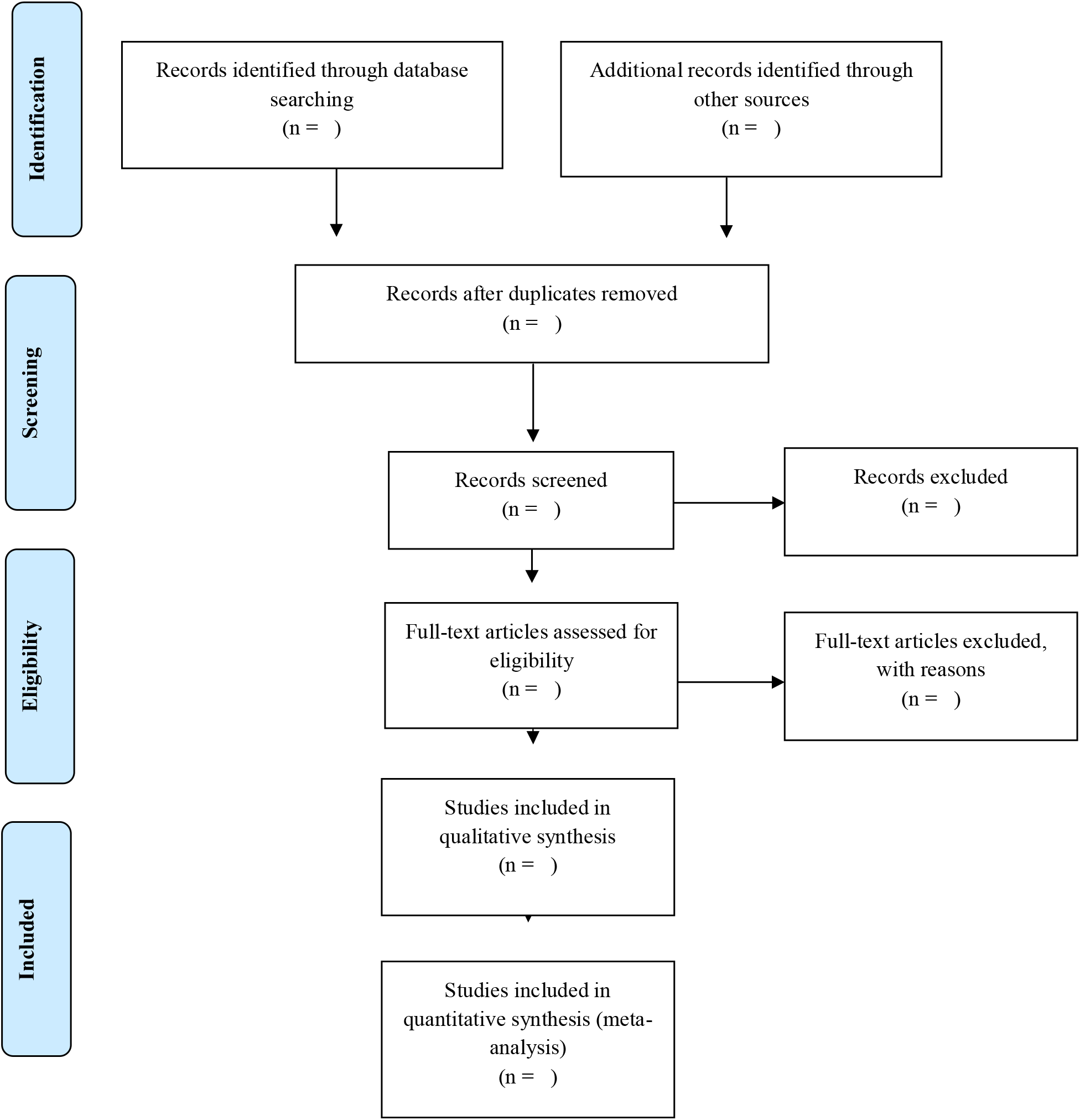
PRISMA Flow Diagram. Source: Adapted from Moher et al.^[53]^

#### Selection of articles

##### Screening by title

After deleting duplicate articles, we will prepare a list of titles of all articles that will be independently screened by a team of two researchers (SGSS and DN), who will independently determine the suitability of articles for the second stage of screening by the abstract. At this stage, all article titles marked as ‘to be excluded’ by both researchers will be removed and all articles marked as ‘to be included’ will be saved as a different file for further screening by the abstract. For articles where the recommendations of both researchers involved in title screening differed from each other, the third reviewer (HCvW) will review the articles and have the final say in either including or excluding an article. Thereafter, a list of all included articles will be prepared for the second screening by reading the abstracts.

##### Screening by abstract

We will read abstracts of all articles retained after the initial articles screening by title will be screened further for relevance to the aim and objectives of the study. The screening of abstracts will be undertaken by the same two researchers (SGSS and DN) involved in the title screening. The process for short listing of articles through abstract screening will be the same as reported under the article screening by title stage.

##### Screening by reading full-text

We will collect the full text of all articles that will be shortlisted at the abstract level screening. A team of two researchers who were involved in the title and abstract screening stages will review the shortlisted articles and will decide whether an article should be included in or excluded from the study. It might be possible that some articles will be found not relevant to the study aim and objectives; hence, such articles will be excluded from the study; while the remaining articles will be included in the last phase of article selection.

##### Selection and inclusion of studies

All of those articles that the two researchers (SGSS and DN) will independently identify as relevant to the study will be included in the systematic literature review and meta-analysis. Any differences between the two researchers will be resolved by the third reviewer (HCvW). Thereafter, full text copies of all articles included in the systematic review will be obtained and read for data extraction as reported below.

### Data extraction process and items

Two independent reviewers (SGSS and DN) will thoroughly read the full text of shortlisted articles. The two reviewers will independently extract information from the shortlisted articles on a predetermined data extraction template (Table 3). Since, avoidance of bias and reduction of errors in the data extractions is imperative in systematic literature reviews and meta-analysis;^[49]^ hence, after the completion of data extraction, the two data extraction forms will be compared and any differences and discrepancies will be reconciled with discussion and in the case of non-agreement between the two reviewers, a third researcher (HCvW) will be involved for arbitration and the final decision.

Since our systematic review will assess the effectiveness; we will therefore extract thorough details of interventions, which is important for the reproducibility of effective interventions.^[54,55]^ We will not extract data at the level of individual patients but the cumulative data from the study, which will be used for the synthesis and inclusion in the meta-analysis as explained below. We will not contact the original researchers with regard to any queries regarding the data reported in their published articles. Identification of multiple reports and publication of the same data could be possible whilst undertaking a systematic review.^[49]^ To deal with the duplicate or multiple reporting of the same data, we will only report the data once that we will find to be comprehensive and reported in a detailed form.

We will extract data on various items as shown in the data extraction template (Table 3), which we have developed a priory in house using an Excel spreadsheet. With regard to the outcomes and measures, our primary outcome of interest in loneliness as measured by any of the loneliness scales as reported earlier.

## DATA SYNTHESIS AND REPORTING RESULTS

There are two options for reporting data in a systematic review on effectiveness statistical synthesis (meta-analysis) and narrative summary (narrative synthesis).^[48]^ We will extract and analyse data at the study level, and we will also pool data from studies that will be sufficiently homogeneous. We will report findings using a summary of the characteristics of included studies and the findings by the type of digital technology intervention, the age, gender and ethnicity of participants, if possible. In addition, we will undertake quantitative synthesis and descriptive/narrative synthesis for quantitative and qualitative studies respectively. Moreover, we will exclude studies with missing values from the meta-analysis.

### Meta-analysis

In the meta-analysis, we will use the effect sizes as measured by common quantitative indicators such as the risk ratio (RR), risk difference (RD) and odds ratio (OR) for the dichotomous outcomes and the weighted mean difference (WMD), and standardised mean difference (SMD) for continuous outcomes.^[49]^ For every study included in our systematic review and meta-analysis, we will calculate the effect size as reported by Masi et al.^[33]^ In addition, we will report a statistical synthesis of our meta-analysis using a statistical summary of RRs, ORs, WMDs and SMDs using the forest plots.^[56]^ For our meta-analysis, we will run the random-effects model as the statistical model^[57,58]^ that is based on the assumption that the true effect size varies between studies and follows a normal distribution around the mean, which is opposite to the fixed effect model based on the assumption that all studies have the same true effect size.^[33]^ We will calculate the Q statistics and p –values for checking the assumption of the homogeneity of effect sizes, and we will determine the T^2^ statistics for estimating the magnitude of the variance of the true effect sizes between studies.^[33]^

### Assessment of research quality, bias and heterogeneity

Two researchers (SGSS and DN) will independently assess the research quality and bias in the selected studies using the validated tools (Table 4). Any discrepancy or disagreement among the reviewers will be resolved by discussion and consensus between them or through arbitration of the third reviewer (HCvW). We will evaluate heterogeneity i.e. variation in study outcomes/effect sizes between studies by the Cochran’s Q test with a significance level of *ρ* < 0.05.^[33,59]^ We will calculate I^2^ statistic^[33,60]^ to determine the proportion of variation in effect size across studies due to heterogeneity considering I^2^ of 25% as low heterogeneity, 50% moderate heterogeneity^[33,60]^ and 75% as a high heterogeneity/variance between studies.^[12,60]^ If the heterogeneity was substantial (I^2^ ≥50%).^[49]^ We would not run the meta-analysis and therefore we will report only a narrative synthesis. For checking the publication bias in the studies included in our systematic review, we will use two methods: graphical method using funnel plots and statistical method using the Egger’s test.^[56,61]^ In assessing the quality of research, we will apply the Grading of Recommendations Assessment, Development and Evaluation (GRADE) approach.^[62]^

**Table 4.**
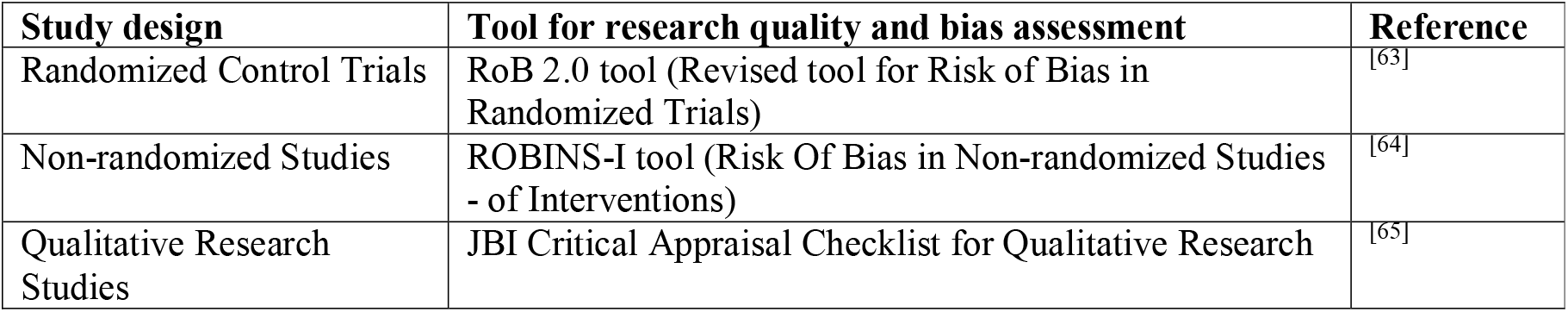
Tool for research quality and bias assessment.

## PATIENT AND PUBLIC INVOLVEMENT

The involvement of patients and public has been suggested in systematic reviews.^[66]^ We could not identify any patient diagnosed with loneliness or a suitable member of public to involve them in the development of this protocol. As such there was no patient and public involvement at the protocol stage in our study, like other published research protocols.^[4,67]^

## ETHICS AND DISSEMINATION

This study is a systematic review and meta-analysis of published research. Hence, we will not seek an ethics approval because we will neither recruit any patients nor analyse data at an individual participant level; however, we will pool data that will be analysed at the study level. We will disseminate our findings through conference papers and presentations and publication of open access articles in peer-reviewed journals.

## DISCUSSION

We believe our research protocol includes a robust research strategy that will enable us to identify and appraise the recent empirical research on digital interventions to reduce loneliness in adult people. Our systematic literature review and meta-analysis will provide the latest evidence on the effectiveness of digital interventions in reducing loneliness among adult people.

We will discuss our findings in relation to our aims and objectives as well as compare and contrast our findings with the current knowledge in the existing body of evidence. We will highlight the contributions, strengths and limitations of our study. In addition, we will discuss the significance, applicability, generalisability and implications of our findings. In addition, we will suggest directions for further research in the domain of loneliness with reference to the application of digital technology interventions.

## CONCLUSION

We anticipate that using this research protocol, we will find primary research using digital interventions to reduce loneliness in adult people. We believe our robust research methodology and analytical strategy will enable us to meet the objectives of our systematic review and meta-analysis. We conclude that appraising the latest empirical research on digital interventions used to reduce loneliness among adults could contribute in informing health and care and public health policy and possibly other stakeholders and private entities (e.g. health insurers) aiming to tackle loneliness in the adult population.

## Data Availability

Not applicable because this is a protocol of a systematic review and meta-analysis.

## ACKNOWLEDGEMENTS

The authors wish to thank Liz Callow of Bodleian Health Care Libraries, Oxford for support in running literature searches.

## AUTHOR CONTRIBUTIONS

All authors were involved in the planning, conception and design of the study. SGSS drafted the manuscript, which was reviewed by DN, HCvW and VK. SGSS undertook preliminary literature searches in the PubMed with the help from Bodleian Health Care Libraries, Oxford. All authors have read, reviewed, contributed in revising and approved the final manuscript. SGSS is the guarantor. VK supervised the study project and arranged funding for the article publication charges.

## FUNDING

This research was funded/supported by the National Institute for Health Research (NIHR) Oxford Biomedical Research Centre. The views expressed are those of the author(s) and not necessarily those of the NHS, the NIHR or the Department of Health.

## COMPETING INTERESTS

Authors declare no competing interests.

## Appendix I PRISMA-P Statement checklist

| Section/topic | # | Checklist item | Reported on page # |
| --- | --- | --- | --- |
| **TITLE** |  |  |  |
| Title | 1 | Identify the report as a systematic review, meta-analysis, or both. | 1 |
| **ABSTRACT** |  |  |  |
| Structured summary | 2 | Provide a structured summary including, as applicable: background; objectives; data sources; study eligibility criteria, participants, and interventions; study appraisal and synthesis methods; results; limitations; conclusions and implications of key findings; systematic review registration number. | 1-2 |
| **INTRODUCTION** |  |  |  |
| Rationale | 3 | Describe the rationale for the review in the context of what is already known. | 2-5 |
| Objectives | 4 | Provide an explicit statement of questions being addressed with reference to participants, interventions, comparisons, outcomes, and study design (PICOS). | 6 |
| **METHODS** |  |  |  |
| Protocol and registration | 5 | Indicate if a review protocol exists, if and where it can be accessed (e.g., Web address), and, if available, provide registration information including registration number. | 1 and 5 |
| Eligibility criteria | 6 | Specify study characteristics (e.g., PICOS, length of follow-up) and report characteristics (e.g., years considered, language, publication status) used as criteria for eligibility, giving rationale. | 6-7 |
| Information sources | 7 | Describe all information sources (e.g., databases with dates of coverage, contact with study authors to identify additional studies) in the search and date last searched. | 7-8 |
| Search | 8 | Present full electronic search strategy for at least one database, including any limits used, such that it could be repeated. | 8 |
| Study selection | 9 | State the process for selecting studies (i.e., screening, eligibility, included in systematic review, and, if applicable, included in the meta-analysis). | 9-10 |
| Data collection process | 10 | Describe method of data extraction from reports (e.g., piloted forms, independently, in duplicate) and any processes for obtaining and confirming data from investigators. | 11 |
| Data items | 11 | List and define all variables for which data were sought (e.g., PICOS, funding sources) and any assumptions and simplifications made. | 6, 11 |
| Risk of bias in individual studies | 12 | Describe methods used for assessing risk of bias of individual studies (including specification of whether this was done at the study or outcome level), and how this information is to be used in any data synthesis. | 12-13 |
| Summary measures | 13 | State the principal summary measures (e.g., risk ratio, difference in means). | 12 |
| Synthesis of results | 14 | Describe the methods of handling data and combining results of studies, if done, including measures of consistency (e.g., $I^2$) for each meta-analysis. | 12 |

Source: Moher et al.<sup>[53]</sup>

## Notes

### Competing Interest Statement

The authors have declared no competing interest.

### Clinical Protocols

http://www.crd.york.ac.uk/PROSPERO/display_record.php?ID=CRD42019131524

### Author Declarations

All relevant ethical guidelines have been followed and any necessary IRB and/or ethics committee approvals have been obtained.

Any clinical trials involved have been registered with an ICMJE-approved registry such as ClinicalTrials.gov and the trial ID is included in the manuscript.

